# Artificial intelligence enabled retinal vasculometry for prediction of circulatory mortality, myocardial infarction and stroke

**DOI:** 10.1101/2022.05.16.22275133

**Authors:** Alicja R Rudnicka, Roshan A Welikala, Sarah A Barman, Paul J Foster, Robert Luben, Shabina A Hayat, Kay-Tee Khaw, Peter H Whincup, David P Strachan, Christopher G Owen, the UK Biobank Eye and Vision Consortium

**Affiliations:** Population Health Research Institute, St George’s, University of London, Cranmer Terrace, London, SW17 0RE, UK; Faculty of Science, Engineering and Computing, Kingston University, Penrhyn Road, Kingston upon Thames, Surrey, KT1 2EE, UK; NIHR Biomedical Research Centre at Moorfields Eye Hospital and UCL Institute of Ophthalmology, London EC1V 2PD, UK; Population and Data Sciences Group, UCL Institute of Ophthalmology, 11-43 Bath Street, London EC1V 9EL, UK; MRC Epidemiology Unit, University of Cambridge, Cambridge, UK; Department of Psychiatry, Cambridge Public Health, University of Cambridge School of Clinical Medicine, Cambridge, UK

## Abstract

**Aims:** We examine whether inclusion of Artificial Intelligence (AI)-enabled retinal vasculometry (RV) improves existing risk algorithms for incident stroke, myocardial infarction (MI) and circulatory mortality.

**Methods:** AI-enabled retinal vessel image analysis processed images from 88,052 UK Biobank (UKB) participants (aged 40-69 years at image capture) and 7,411 EPIC-Norfolk participants (aged 48-92). Retinal arteriolar and venular width, tortuosity and area were extracted. Prediction models were developed in UKB using multivariable Cox proportional hazards regression for circulatory mortality, incident stroke and MI, and externally validated in EPIC-Norfolk. Model performance was assessed using optimism adjusted calibration, C- and R^2^ statistics. Performance of Framingham risk scores (FRS) for incident stroke and incident MI, with addition of RV to FRS, were compared with a simpler model based on RV, age, smoking status and medical history (antihypertensive/cholesterol lowering medication, diabetes, prevalent stroke/MI).

**Results:** UKB prognostic models were developed on 65,144 participants (mean age 56.8; median follow-up 7.7 years) and validated in 5,862 EPIC-Norfolk participants (67.6, 9.1 years respectively). Prediction models for circulatory mortality in men and women had optimism adjusted C- and R^2^ statistics between 0.75-0.77 and 0.33-0.44 respectively. For incident stroke and MI, addition of RV to FRS did not improve model performance in either cohort. However, the simpler RV model performed equally or better than FRS.

**Conclusion:** RV offers an alternative predictive biomarker to traditional risk-scores for vascular health, without the need for blood sampling or blood pressure measurement. Further work is needed to examine RV in population screening to triage individuals at high-risk. (250 words)

**HIGHLIGHTS:** 

**What is already known on this topic:**

- Population screening for MI and stroke using risk prediction tools exist but have limited uptake; risk scores for circulator mortality do not exist.

**What this study adds:**

- Risk models developed in UK Biobank (validated in EPIC-Norfolk) using Artificial Intelligence (AI)-enabled retinal vasculometry (RV), age, history of cardiovascular disease, use of hypertensive medication and smoking yielded high predictive test performance for circulatory mortality.
- Risk scores for MI and stroke performed similarly to established risk scores.

**How this study might affect research, practice or policy:**

- AI-enabled RV extraction offers a non-invasive prognostic biomarker of vascular health that does not require blood sampling or blood pressure measurement, and potentially has greater community reach to identify individuals at medium-high risk requiring further clinical assessment.

**SYNOPSIS/PRECIS:** Risk models developed in UK Biobank (validated in EPIC-Norfolk) using Artificial Intelligence enabled retinal vasculometry indices, age, history of cardiovascular disease, use of hypertensive medication and smoking yielded high predictive test performance for circulatory mortality. Risk scores for MI and stroke performed similarly to established risk scores.

## INTRODUCTION

Circulatory mortality, including cardiovascular disease (CVD), coronary heart disease (CHD), heart failure and stroke, is a major cause of morbidity and mortality worldwide.^1,2^ A large number of risk algorithms exist to predict CVD,^3^ and the addition of fixed and modifiable risk factor phenotypes have been evaluated, but have so far shown little improvement in CVD prediction.^4-6^ Machine learning techniques incorporating 473 potential risk factors for the prediction CVD in the UK Biobank cohort yielded areas-under-the-curve (AUC) from receiver operating characteristic curve of 0.774, compared with AUC of 0.724 for Framingham risk scores.^7^ Other CVD risk scores, using different CVD outcome definitions, have already been evaluated in UK Biobank including the European Systemic Coronary Risk Evaluation (SCORE)^8^, QRISK3^9^ and American College of Cardiology/American Heart Association^10^ risk score with C-statistic values of 0.775, 0.739 and 0.736 respectively.^6^

Examination of retinal blood vessels (arterioles and venules) may offer a microvascular phenotype more indicative of the presence of early circulatory related disease processes, providing a non-invasive window on the circulatory system. Narrow retinal arterioles show a clear association with higher blood pressure, hypertension and with incident CVD.^11^ Arteriolar vessel width narrowing and venular widening may be important for mortality, stroke ^11^ and CHD incidence^12^, but there are inconsistencies in the literature,^13 14^ such as retinal vessel associations with CVD risk in women but not in men.^11,15^ Other features of retinal vasculometry (RV), such as vessel tortuosity, may offer more discerning markers of vascular status but remain little studied at scale.^16,17^ Unfortunately, machine learning approaches do not currently clarify which features of RV are important, although they may do in future.

We developed a fully automated AI-enabled system (QUARTZ) for examining the retinal vascular tree, which overcomes many of the difficulties of earlier approaches, allowing detailed vasculometry quantification in large population studies.^18-20^ In the subset of UK Biobank who underwent retinal imaging,^21^ and in the EPIC-Norfolk^16^ cohorts, we examine detailed characterisation of RV as a non-invasive maker of vascular health in relation to circulatory mortality prediction. In addition, we provide findings for Framingham risk scores for stroke,^22^ and MI^23^ in the same subset that underwent retinal imaging, and assess the incremental value of adding RV to Framingham risk scores for incident stroke and MI.

## MATERIALS AND METHODS

### UK Biobank

is a prospective cohort study for which baseline biomedical and physical assessments were carried out 2006-2013, in 502,682 adults aged 40-69 years recruited from 22 UK centres.^24^ Ocular assessments occurred during the latter phase (2009-2013; 7 centres) and included visual acuity, autorefraction, digital fundus photography with the Topcon 3D-OCT 1000 Mark 2.^21^ Non-mydriatic 45° digital colour images, centred on the fovea were available for 88,052 participants.

### EPIC-Norfolk

was the UK component of the European Prospective Investigation into Cancer (EPIC) study.^25,26^ Here we focus on data from the 3^rd^ clinical follow-up (2004-2011)^27^ on 8,603 participants aged 48-92 years who underwent a biomedical and eye examination similar to that of UK Biobank (see Supplementary Material for further details).^16^

### Health outcomes

The primary outcome was circulatory mortality as defined using International Classification of Diseases (ICD) (ICD-10 codes I00-I99 and ICD9 390-459) coded death registry data from the Office for National Statistics and the Health and Social Care Information Centre (now NHS Digital) for England and Wales, and the Information Services Department for Scotland, provided information on date and cause(s) of death to 31^st^ January 2018 for UK Biobank and 31^st^ March 2018 for EPIC-Norfolk. Incident MI and stroke events after retinal image capture were based on medical records linkage with hospital diagnoses of non-fatal events, supplemented with participant health and lifestyle questionnaire data from repeat surveys in UK Biobank and EPIC-Norfolk (2012-2018). ICD-10 codes I21-I25 (or ICD-9 codes 410, 411,412 429.79) were used for fatal and non-fatal MI; and ICD-10 codes I60,61,63,64 (or ICD-9 codes 430, 431,434,436) for ischaemic and haemorrhagic stroke (see Algorithmically - define health outcomes https://www.ukbiobank.ac.uk/enable-your-research/about-our-data/health-related-outcomes-data).

### AI-enabled retinal image processing

A validated, fully automated AI-enabled system (QUARTZ)^18-20^ extracted thousands of measures of retinal vessel width, tortuosity and area from the whole retinal image. Supervised machine learning techniques were used within QUARTZ; with a support vector machine used to create an image quality score^18^ and deep learning was used to develop an algorithm to distinguish between arterioles and venules.^19^ QUARTZ measures of width (microns^28^), total vessel area (mm^2^), tortuosity (arbitrary units),^16,29^ and variance of widths along a vessel segment, were averaged for each image (weighted by the length of each vessel segment), separately for arterioles and venules. Person level averages were obtained by averaging across right and left eyes.

### Statistical Analysis

Statistical analyses were carried out using STATA software (version 16, StataCorp LP, College Station, TX). Retinal vessel widths and area showed normal distributions, tortuosity required log-transformation and within-vessel-width-variance required inverse square-root transformation to normalize distributions. Models were developed in UK Biobank for men and women separately throughout, and externally validated in EPIC-Norfolk. We hypothesized that retinal vessel characteristics in relation to circulatory mortality might be modified by age, smoking status, presence of CVD/diabetes and use of BP lowering medications. Hence, two-way interactions between RV and age, smoking status and self-reported use of blood pressure medication, prevalent diabetes and CVD were first examined in mutually adjusted Cox proportional hazard^30^ models for circulatory mortality. Interaction terms with p values <0.2 were then included along with main effects in Cox regressions models using backward elimination (p value set to 0.1). Bootstrapping with 100 replications was used for internal validation to adjust model performance measures for optimism, including Harrel’s C-statistic for discrimination, R^2^ statistic (representing a measure of explained variation)^31^ and calibration slope (where a slope of 1.0 is ideal).^32^ The original beta coefficients adjusted for shrinkage were applied to EPIC-Norfolk to estimate C-statistic, R^2^ and calibration slopes. Model performance was graphically assessed from plots of the observed probability of event at 5 years by deciles of predicted risk at 5 years in UK Biobank and by octiles in EPIC-Norfolk.

Framingham risk scores (FRS) for incident fatal and non-fatal stroke^22^ and MI^23^ were applied to UK Biobank and EPIC-Norfolk cohorts and recalibrated to baseline survival function within each cohort according to the 5 year survival rates. Following FRS criteria, participants reporting use of cholesterol lowering medications, diabetes or missing data on total or HDL cholesterol were excluded from all MI analyses.^23^ FRS models were also extended to include RV. Alternative models for incident fatal and non-fatal stroke and MI using age, smoking status, medical history (self-reported history of heart attack, stroke or diabetes and use of blood pressure lowering medications) and RV only were developed in UK Biobank following the same approach as for circulatory mortality. A medical history of MI did not preclude inclusion in models for incident stroke events, and vice-versa.

Sensitivity analyses restricted model development and validation to white ethnicity. Using EPIC-Norfolk, external validation was extended to a broader spectrum of incident cerebrovascular disease (ICD10 I60-69; ICD 9 430-438) and incident ischaemic heart disease (ICD10 I20-I25; ICD9 410-414). We follow TRIPOD guidelines for reporting of model development and validation.^33^

## RESULTS

Table 1 shows for UK Biobank mean age at baseline was 56.8 years with median duration of follow-up 7.7 years after retinal image capture (maximum 8.2 years), and for EPIC-Norfolk, mean age was older (67.6 years) and median follow-up 9.1 years (maximum 12.4 years). Figure 1 is a visual representation of retinal image analysis using the QUARTZ software. Figure S1 shows the number of UK Biobank and EPIC-Norfolk participants and events available for circulatory mortality, incident stroke and incident MI analyses.

**Table 1.**
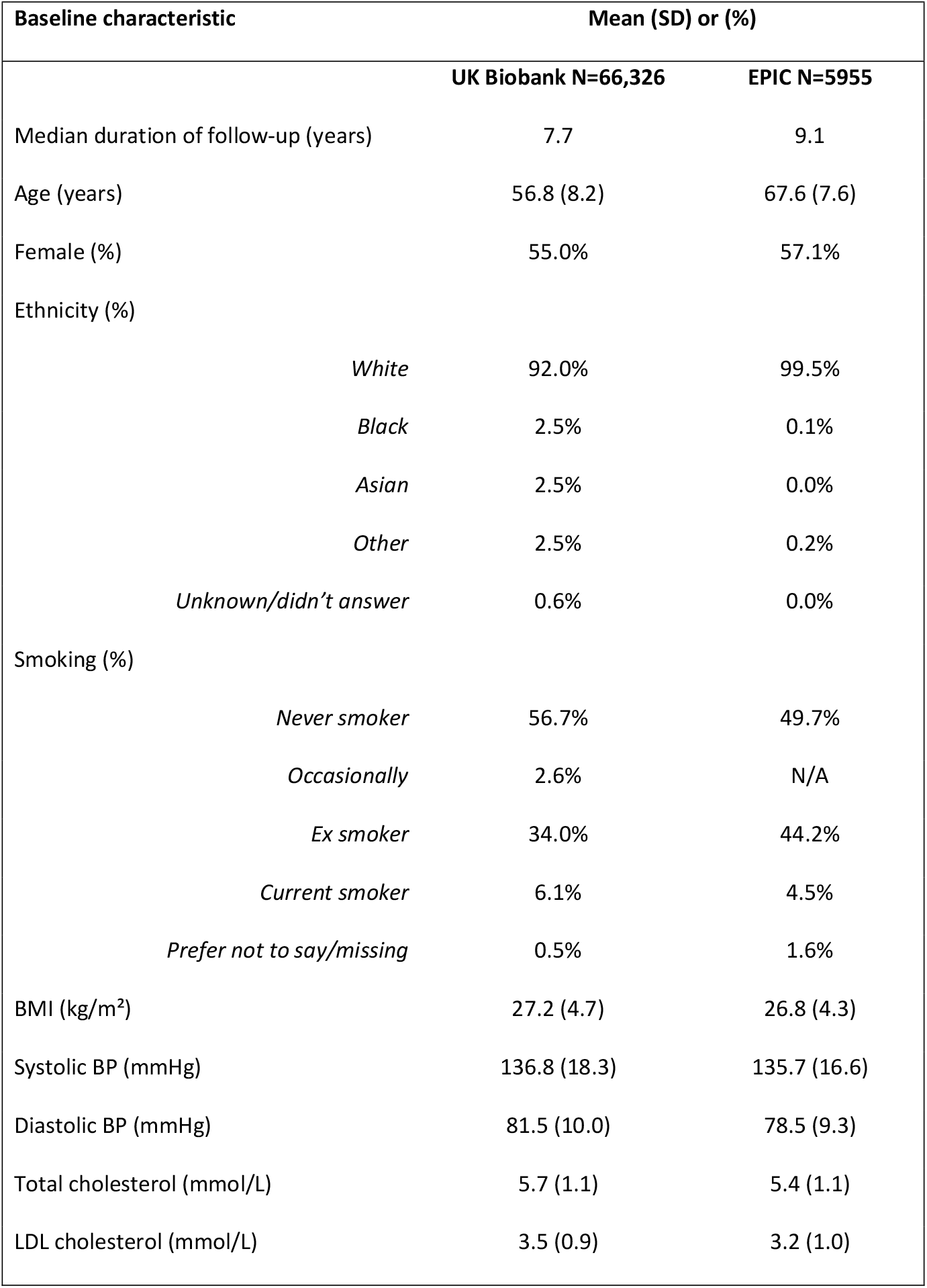
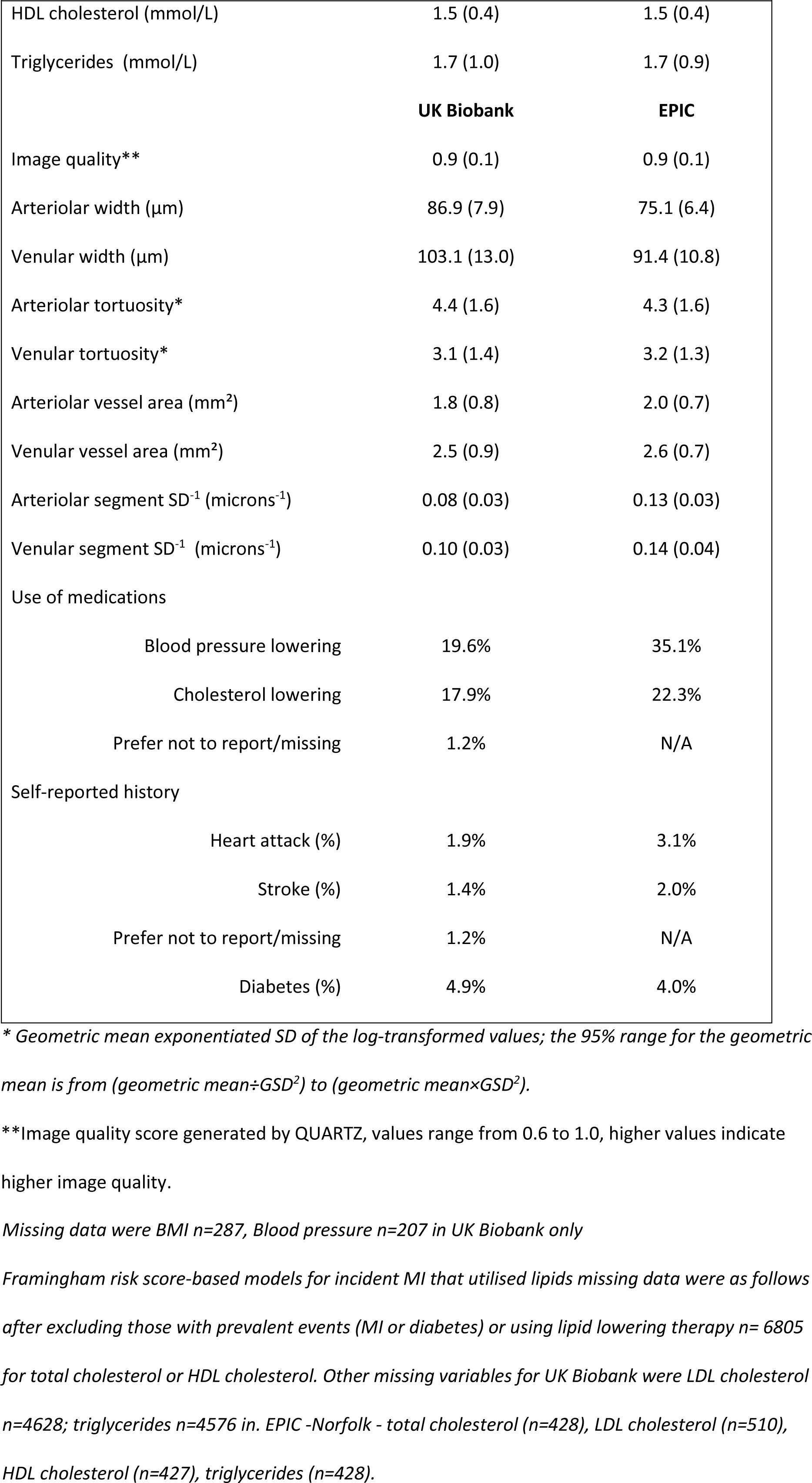
Clinical characteristics at baseline eye assessment in UK Biobank (2009-2013) and from the third health check phase for EPIC-Norfolk (2004-2011). Values are mean (SD) or (%)

**Figure 1:**
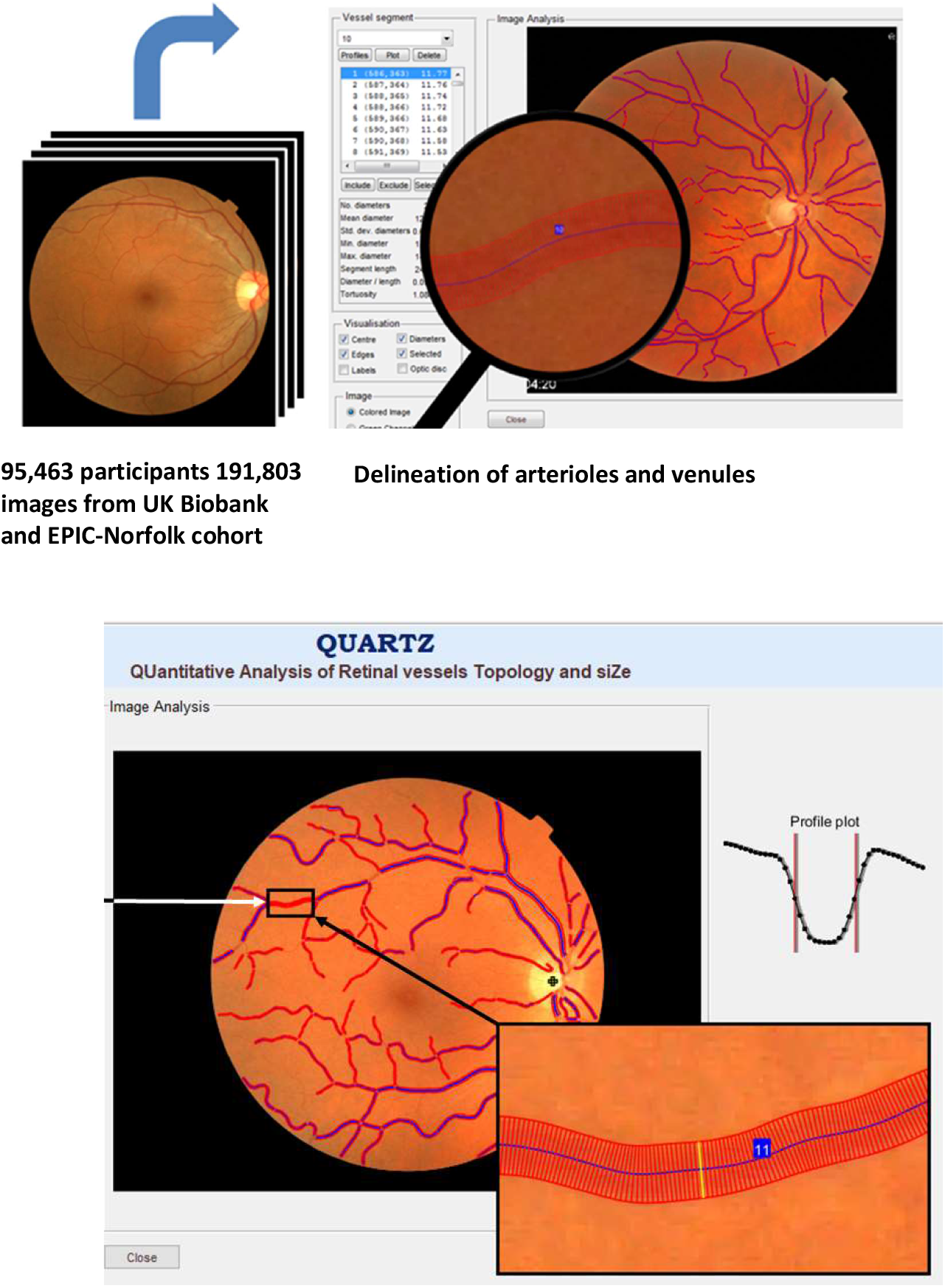
Fully automated retinal image processing of the vascular tree using AI-enabled QUARTZ software **Retinal vasculometry measures include vessel segment width, tortuosity and within vessel segment variation in width for arterioles and for venules separately**

### Circulatory mortality

64,144 UK Biobank participants with 327 circulatory deaths and 5862 EPIC-Norfolk participants with 201 circulatory deaths were included. In men, arteriolar and venular width, tortuosity and width-variance were identified as statistically significant predictors of circulatory mortality. In women, arteriolar and venular area and width, venular tortuosity and venular width-variation contributed to risk prediction. RV effects on circulatory mortality were modified by smoking status, blood pressure medications and history of MI. In men and women, optimism adjusted C- (0.75 to 0.77) and R^2^ (0.33 to 0.44) statistics in UK Biobank and EPIC-Norfolk, were reasonably high (Table 2, Table S1 for full model diagnostics; Tables S2 and S3 for regression coefficients). In UK Biobank men, predicted risks were closely aligned with observed risks. A similar picture emerged for EPIC-Norfolk men cohort, with about double the risk of circulatory mortality, half the numbers of events and being about a decade older at retinal image capture had (Figure 2). UK Biobank women showed a wide separation of risk groups and close alignment of predicted and observed risks even, at low risks (<0.5%). Calibration plots for EPIC-Norfolk women were less clear due to the lower number of events available, hence 95% CI around predictions were wider (Figure 2).

**Table 2.** Optimism adjusted model performance (95% confidence intervals) for prediction of circulatory mortality, incident stroke and myocardial infarction models developed in UK Biobank cohort (2009-2018) with external validation in EPIC-Norfolk cohort (2004-2018). Framingham risk scores (FRS) for incident stroke and myocardial infarction are also presented.

| Model | UK Biobank Men | UK Biobank women | EPIC-Norfolk Men | EPIC-Norfolk women |
| --- | --- | --- | --- | --- |
| **CIRCULATORY MORTALITY (number of events/sample size)** |  |  |  |  |
| **Age, smoking, medical history + RV** | (227/29,257) | (100/35,887) | (114/2,516) | (87/3,346) |
| Calibration Slope | 0.913 (0.800, 1.026) | 0.857 (0.732, 0.982) | 1.084 (0.888, 1.279) | 0.872 (0.674, 1.070) |
| C-statistic | 0.749 (0.720, 0.779) | 0.763 (0.717, 0.810) | 0.774 (0.732, 0.815) | 0.748 (0.692, 0.805) |
| R<sup>2</sup> | 0.369 (0.310, 0.427) | 0.443 (0.369, 0.518) | 0.392 (0.302, 0.482) | 0.333 (0.228, 0.438) |
| **STROKE (number of events/sample size)** |  |  |  |  |
| **FRS for stroke** | (245/28,573) | (201/35,266) | (98/2,432) | (113/3,276) |
| Calibration Slope | 0.908 (0.769, 1.047) | 0.919 (0.764, 1.074) | 0.819 (0.552, 1.087) | 0.943 (0.734, 1.152) |
| C-statistic | 0.736 (0.706, 0.766) | 0.736 (0.702, 0.770) | 0.682 (0.629, 0.735) | 0.732 (0.682, 0.781) |
| R<sup>2</sup> | 0.295 (0.233, 0.358) | 0.310 (0.240, 0.379) | 0.199 (0.098, 0.300) | 0.309 (0.215, 0.402) |
| **Age, smoking, medical history + RV** |  |  |  |  |
| Calibration Slope | 0.896 (0.767, 1.025) | 0.860 (0.729, 0.991) | 0.808 (0.571, 1.045) | 0.780 (0.603, 0.958) |
| C-statistic | 0.729 (0.699, 0.759) | 0.753 (0.721, 0.784) | 0.691 (0.637, 0.746) | 0.714 (0.660, 0.768) |
| R<sup>2</sup> | 0.315 (0.256, 0.375) | 0.352 (0.289, 0.416) | 0.213 (0.113, 0.314) | 0.274 (0.179, 0.369) |
| **MYOCARDIAL INFARCTION (number of events/sample size)** |  |  |  |  |
| **FRS for confirmed MI** | (275/19,150) | (118/26,584) | (166/1,622) | (99/2,440) |
| Calibration Slope | 1.216 (0.994, 1.439) | 1.036 (0.813, 1.260) | 1.567 (1.210, 1.924) | 0.834 (0.583, 1.085) |
| C-statistic | 0.706 (0.678, 0.734) | 0.758 (0.718, 0.798) | 0.689 (0.650, 0.728) | 0.688 (0.640, 0.737) |
| R<sup>2</sup> | 0.235 (0.175, 0.295) | 0.345 (0.256, 0.433) | 0.233 (0.153, 0.312) | 0.208 (0.109, 0.308) |
| **Age, smoking, medical history + RV** |  |  |  |  |
| Calibration Slope | 0.836 (0.673, 0.999) | 0.803 (0.590, 1.016) | 0.905 (0.655, 1.156) | 0.786 (0.517, 1.054) |
| C-statistic | 0.675 (0.647, 0.703) | 0.709 (0.669, 0.749) | 0.641 (0.598, 0.683) | 0.650 (0.593, 0.707) |
| R<sup>2</sup> | 0.178 (0.118, 0.238) | 0.226 (0.136, 0.316) | 0.150 (0.077, 0.224) | 0.162 (0.067, 0.256) |

FRS= Framingham risk score. RV = retinal vasculometry
Estimates for Calibration slope, C-statistic and R<sup>2</sup> values are given with bootstrapped 95% confidence interval in parenthesis

**Figure 2:**
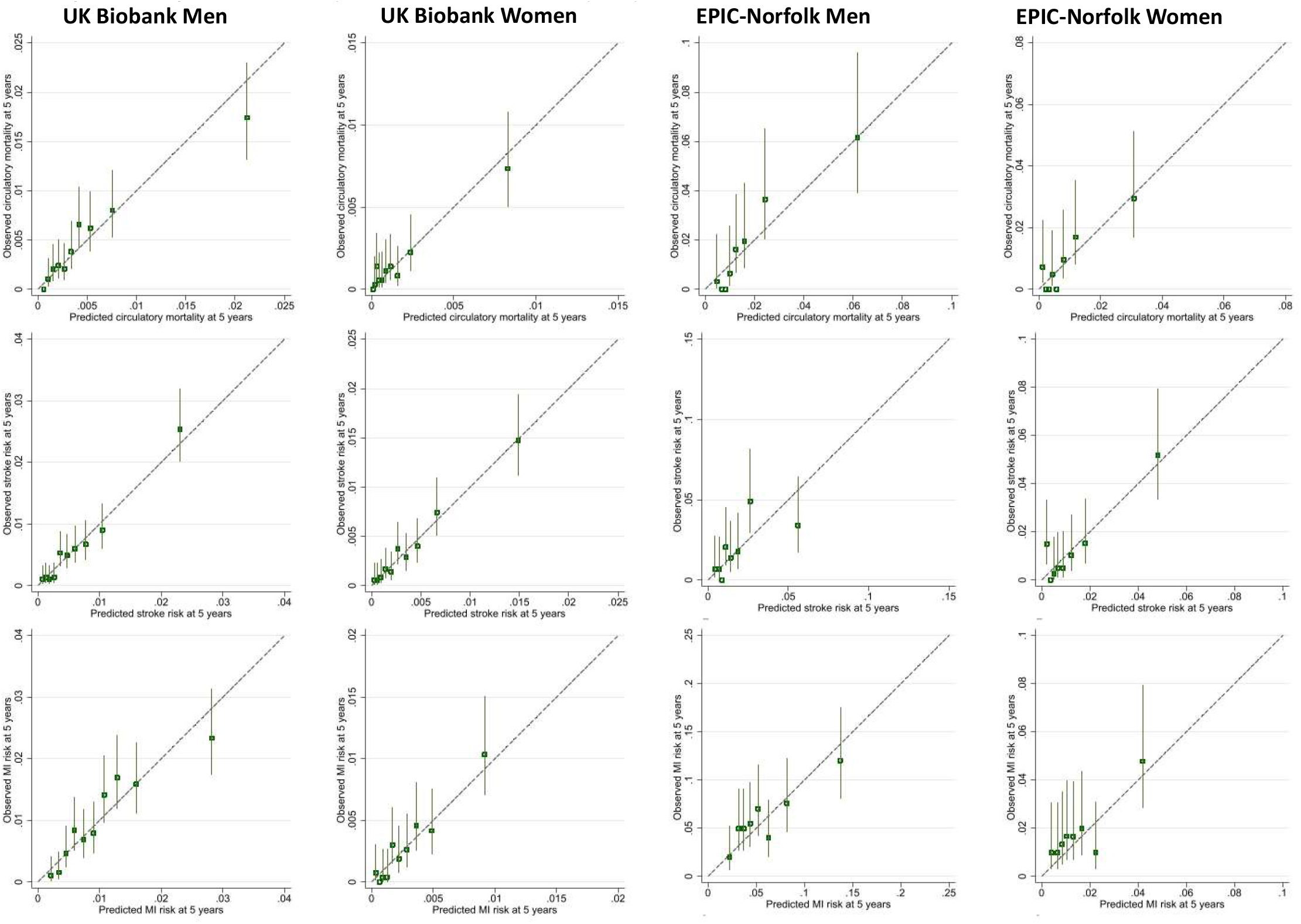
Observed risk of outcome at 5 years by deciles of predicted risk in UK Biobank and eighths of predicted risk in EPIC-Norfolk. Prediction models based on age, smoking, medical history with retinal vasculometry only. **Figure 2 footnote**: Top row: Circulatory mortality Middle row: Incident stroke Bottom row Incident MI Vertical lines around symbols are 95% confidence intervals. Dotted line represents perfect calibration. The scale of the vertical and horizontal axes is a probability e.g., 0.1 equates to a 10% risk of event by 5 years. Circulatory mortality codes: ICD10: I00-I69, ICD9: 390-359 Incident stroke codes: ICD10: I60,I61,I63,I64, ICD9: 430,431,434,436 Incident MI codes: ICD10: I21-I25, ICD9: 410,411,412,429.79

### Incident stroke

63,839 UK Biobank participants with 446 incident strokes and 5,708 EPIC-Norfolk participants with 211 incident stroke events after retinal image capture were included (Figure S1). In UK-Biobank, FRS C-statistic was 0.74 in men and 0.74 in women (Table 2) with lower values in EPIC-Norfolk; approximately one third of the variation in stroke-risk incidence was explained by R^2^ (less so in EPIC-Norfolk men). Observed risks were more aligned with predicted risks in men than in women (Figure S2). Addition of RV to FRS did not improve model performance statistics overall (Table S4; Figure S2).

Models based on age, smoking status, medical history and RV showed similar performance to FRS with C-statistic of 0.73 in men and 0.75 in women and marginally improved R^2^ values in UK Biobank; (Table 2; full model diagnostics Tables S4). As for FRS, performance metrics were lower in EPIC-Norfolk. Multivariable models (Tables S2-S3) showed venular and arteriolar tortuosity and width were predictors of stroke in men and women and additionally venular/arteriolar area in women with some modification by smoking status, blood pressure medications and history of MI. Calibration plots showed risk predictions closer to the 45-degree line particularly at lower levels of predicted risk in women (Figure S2).

### Incident MI

45,734 UK Biobank participants with 393 incident MI and 4,062 in EPIC-Norfolk with 265 incident MI after retinal image capture were included (Figure S1). In UK Biobank, FRS C-statistics were 0.71 in men and 0.76 in women with approximately one quarter (24%) of the variation in MI risk explained by FRS in men and 35% in women (Table 2). In EPIC-Norfolk, with approximately 5x the risk of MI, performance statistics were lower. Calibration plots for FRS showed better alignment of observed and predicted risks in men compared with women (Figure S3). Addition of RV to FRS did not improve model performance overall (Table S5; Figure S3). Compared to FRS alone a simpler model based on age, smoking status, medical history and RV performed marginally less well in men and women in both cohorts. (Tables 2 and S5; Figure S3). Multivariable models for MI using RV (Table S2-S3) showed arteriolar and venular width, venular width variability and arteriolar area were predictors in men, whereas for women venular tortuosity, venular/arteriolar area and venular width variability were predictors. RV effects were modified by smoking status.

### Cases in top quintile of risk scores

For circulatory mortality models based on age, smoking status medical history and RV captured between 52-65% of cases of circulatory mortality in the top quintile of the risk score distribution (Table 3). For incident stroke, RV based models compared with FRS captured about 5% more cases in UK Biobank men and 8% more cases in UK Biobank women and 3% more EPIC-Norfolk men in the top quintile of risk scores (Table 3) but 1.8% fewer EPIC-Norfolk women. However, for MI, FRS captured more cases of MI in the top quintile of risk. Considering stroke and MI scores combined, the simpler RV models captured more cases in the top quintile than FRS for UK Biobank men and women, and similar proportions in EPIC-Norfolk men and women.

**Table 3.** Percentage of circulatory mortality, incident stroke and incident MI events (after retinal image capture) in top quintile of risk score distributions for UK Biobank and EPIC-Norfolk.

| Model | UK Biobank |  | UK Biobank |  | EPIC-Norfolk |  | EPIC-Norfolk |  |
| --- | --- | --- | --- | --- | --- | --- | --- | --- |
|  | Men |  | Women |  | Men |  | Women |  |
|  | Number, % of all circulatory mortality in top quintile of circulatory mortality risk score distribution |  |  |  |  |  |  |  |
| Age, smoking, medical history + RV | 126 | 55.5% | 65 | 65.0% | 63 | 55.3% | 45 | 51.7% |
|  | Number, % of all incident stroke in top quintile of stroke risk score distribution |  |  |  |  |  |  |  |
| FRS stroke | 115 | 46.9% | 100 | 49.8% | 37 | 37.8% | 55 | 48.7% |
| Age, smoking, medical history + RV | 133 | 54.3% | 114 | 56.7% | 40 | 40.8% | 53 | 46.9% |
|  | Number, % of all incident MI in top quintile of MI risk score distribution |  |  |  |  |  |  |  |
| FRS confirmed MI | 116 | 42.2% | 59 | 50.0% | 68 | 41.0% | 37 | 37.4% |
| Age, smoking, medical history + RV | 109 | 39.6% | 58 | 49.2% | 65 | 39.2% | 33 | 33.3% |
|  | Number, % of all incident stroke or MI in top quintile of stroke or MI risk score distribution |  |  |  |  |  |  |  |
|  | Men |  | Women |  | Men |  | Women |  |
| FRS confirmed MI or FRS stroke | 259 | 49.8% | 181 | 56.7% | 120 | 45.5% | 108 | 50.9% |
| Age, smoking, medical history + RV | 264 | 50.8% | 190 | 59.6% | 119 | 45.5% | 104 | 49.9% |

FRS= Framingham risk score RV = retinal vasculometry

### Sensitivity analyses

Restricting model development and validation to those of white ethnicity did not materially alter model performance for any of the models presented. FRS and all RV models for stroke showed systematically improved external validation for outcomes based on inclusion of all incident cerebrovascular disease in EPIC-Norfolk (Table S4 far right-hand column and Figure S4), especially in women. In contrast, for *all* incident ischaemic heart disease in EPIC-Norfolk, performance of FRS and RV models remained remarkably unchanged in men but marginally improved in women (Table S5 far right-hand column and Figure S5).

## DISCUSSION

This study compares risk predictions using AI-enabled RV with established CVD risk-algorithms. To the best of our knowledge it represents the largest population-based study of RV. Importantly, external validation of the prediction models was carried out in a separate large cohort, which is uncommon in this field. Our automated AI-enabled system extracts the retinal vascular tree over the entire retinal image (Figure 1), distinguishes between arterioles and venules and provides measures of tortuosity, width-variance and area, in addition to vessel width. Risk models showed that all RV components contributed to risk prediction. Adding RV to FRS resulted in marginal changes in the prediction of stroke or MI. However, a simpler non-invasive risk score based on age, sex, smoking status, medical history and RV yielded comparable performance to FRS, without the need for blood sampling or blood pressure measurement. Prediction of circulatory mortality using age, sex, smoking status, medical history and RV has not been reported previously, and yielded the highest model performance in terms of C-R^2^ statistics and agreement between observed and predicted risks, even at lower levels of risk, in both the internal and external validation cohorts.

### Comparisons with other studies

Prospective associations have been largely based on retinal vessel width with mortality,^13^ incident stroke,^13 13^ and with CHD (in women, not men),^11,34^ from restricted measurement areas of the retina.^11,35^ Measurements are often not automated, requiring operator involvement, which limits application to large populations. In agreement with others, our models show that both arteriolar and venular vasculometry contribute to risk prediction,^11,36^ and this aligns with our previous work.^16,29,37^ Seidelmann reported that narrower central retinal artery and wider central retinal vein equivalent dimensions offered significant additional information to equations for incident atherosclerotic CVD risk,^11^ especially in women, but C-statistics were modest (between 0.55-0.57) compared with the much higher levels in the current study (i.e., between 0.70-0.77). Our RV models generally performed better in women and may indicate that microvascular dysfunction contributes more to CHD pathogenesis in women than in men, as they have smaller coronary arteries exhibiting more diffuse ‘non-obstructive’ atherosclerosis,^38^ with a larger burden of coronary microvascular disease,^39^ leading to higher morbidity and mortality.^40^ A recent study using the UK Biobank data source in fewer participants (54,813 vs 65,144 in this study), showed that retinal vessel density and fractal dimensions (extracted from the entire image after deep learning vessel segmentation without distinction between arterioles and venules) were associated with other health outcomes, including overall mortality, hypertension and congestive heart failure, but did not report on risk predictionperformance.^41^ Moreover, there was no consistent evidence of associations with incident circulatory disease, and cerebrovascular disease and associations with incident MI were null.^41^ Another study in a sub-set of UK Biobank participants (n=5,663) with both retinal and cardiovascular magnetic resonance imaging used deep learning /AI approaches to estimate structural cardiac indices as intermediaries for predicting MI.^42^ However, given their approach, specific retinal features of importance remain unclear.

European Systematic Coronary Risk Evaluation (SCORE) CVD risk score^8^, QRISK3 risk score^9^ and the American College of Cardiology/American Heart Association CVD risk score algorithms have already been evaluated in UK Biobank. The published C-statistics for these three risk scores were 0.77, 0.74 and 0.74 respectively, with 95%CI that overlap with values for the simpler RV model presented in this study. However, the novel C-statistics for circulatory mortality reported in this study are higher. Our approach of focussing on the retinal microvasculature as a key prognostic marker of incident cardiovascular outcomes and circulatory mortality is supported by saliency maps presented in a study using end-to-end AI of retinal images to estimate the extent of coronary artery calcium scores in cross-sectional associations,^43^ with C-statistics for incident CVD varying between 0.68 to 0.76. Our model using RV together with easily attainable data including age, smoking status, sex and a brief medical history, is simple, non-invasive and exhibits performance that is comparable, or even better than, current risk algorithms, including end-to-end AI approaches.

### Strengths and limitations

Model development in UK Biobank provided a large sample size and number of prospective events. QUARTZ successfully processed a high percentage (∼80%) of retinal images captured by non-experts providing ‘vasculomic’ indices of vascular health. External validation in an older higher risk cohort (EPIC-Norfolk) replicated the findings, and models were also robust to inclusion of a wider spectrum of cerebrovascular and ischaemic heart disease events.

UK Biobank and EPIC-Norfolk are ‘healthy’ cohorts with relatively low event rates compared with other geographically similar middle-aged cohorts.^44^ Prevalence of current smoking was very low in UK Biobank (6%) and limited the ability to examine interactions with RV. Although we did not find limiting the analysis to those of white ethnicity materially altered the results, the proportion of non-white participants in UK Biobank is low. RV may relate to microvascular endothelial function elsewhere in the body and may underpin the causal pathways behind prognostic models, which may differ with ethnicity. Confirmation of model performance in other cohorts with higher CVD rates and in different (especially non-white) ethnic groups would be informative.

### Implications and Conclusions

Retinal imaging is established within clinic and hospital eye care and in optometric practices in the US and UK. AI-enabled vasculometry risk prediction is fully automated, low cost, non-invasive and has the potential for reaching a higher proportion of the population in the community because of ‘high street’ availability and because blood sampling or sphygmomanometry are not needed. RV is a microvascular marker, hence offers better prediction for circulatory mortality and stroke compared with MI which is more macrovascular, expect perhaps in women. In the general population it could be used as a non-contact form of systemic vascular health check, to triage those at medium-high risk of circulatory mortality for further clinical risk assessment and appropriate intervention. Having a further low cost, accessible, non-invasive screening test in the community to encourage clinical risk assessment uptake in the community (in addition to current screening approaches), would help to prolong disease-free status in an ever-ageing population with increasing co-morbidities, and assist with minimising healthcare costs associated with lifelong vascular diseases.

## Data Availability

The data reported in this article are available via application to the UK Biobank to other researchers for purposes of reproducing the results or replicating the procedure.

## CONTRIBUTION STATEMENT

All Authors contributed to this manuscript. ARR, CGO, PHW, DPS, SAB, PJF designed the present study and raised funding. ARR, CGO, RAW, SAB, RL, SAH, KTK, PJF collected data for the study and undertook data management. ARR, RAW, SAB analysed the data. ARR and CGO wrote the first draft of the report, which all authors contributed to and critically appraised. ARR and CGO are responsible for data integrity and will act as guarantors.

## ETHICS, GOVERNANCE AND CONSENT

The UK Biobank and EPIC-Norfolk studies was carried out following the principles of the Declaration of Helsinki and the Research Governance Framework for Health and Social Care. The UK Biobank study was approved by the North West Multi-Centre Research Ethics Committee (11/NW/03820). All participants gave written, informed consent. The EPIC-Norfolk study was approved by the Norfolk Local Research Ethics Committee (05/Q0101/191) and East Norfolk and Waveney NHS Research Governance Committee (2005EC07L). All participants gave written, informed consent.

## ACKNOWLEDGEMENTS

The authors wish to thank Nicola Kimber and Abigail Britton from the MRC Epidemiology Unit, University of Cambridge, for continued support with EPIC-Norfolk data preparation and access.

## UK BIOBANK EYES AND VISION CONSORTIUM

Prof Naomi ALLEN, University of Oxford

Prof Tariq ASLAM, The University of Manchester

Dr Denize ATAN, University of Bristol

Prof Sarah BARMAN, Kingston University

Prof Jenny BARRETT, University of Leeds

Prof Paul BISHOP, The University of Manchester

Prof Graeme BLACK, The University of Manchester

Dr Tasanee BRAITHWAITE, St Thomas’ Hospital

Dr Roxana CARARE, University of Southampton

Prof Usha CHAKRAVARTHY, Queen’s University Belfast

Dr Michelle CHAN, Moorfields Eye Hospital

Dr Sharon CHUA, UCL Institute of Ophthalmology

Dr Alexander DAY, Moorfields Eye Hospital

Dr Parul DESAI, Moorfields Eye Hospital

Prof Bal DHILLON, University of Edinburgh

Prof Andrew DICK, University of Bristol

Dr Alexander DONEY, University of Dundee

Dr Cathy EGAN, Moorfields Eye Hospital

Prof Sarah ENNIS, University of Southampton

Prof Paul FOSTER, UCL Institute of Ophthalmology

Dr Marcus FRUTTIGER, UCL Institute of Ophthalmology

Dr John GALLACHER, University of Oxford

Prof David (Ted) GARWAY-HEATH, UCL Institute of Ophthalmology

Dr Jane GIBSON, University of Southampton

Prof Jeremy GUGGENHEIM, Cardiff University

Prof Chris HAMMOND, King’s College London

Prof Alison HARDCASTLE, UCL Institute of Ophthalmology

Prof Simon HARDING, University of Liverpool

Dr Ruth HOGG, Queen’s University Belfast

Dr Pirro HYSI, King’s College London

Prof Pearse KEANE, UCL Institute of Ophthalmology

Prof Sir Peng Tee KHAW, UCL Institute of Ophthalmology

Dr Anthony KHAWAJA, Moorfields Eye Hospital

Mr Gerassimos LASCARATOS, Moorfields Eye Hospital

Dr Thomas LITTLEJOHNS, University of Oxford

Prof Andrew LOTERY, University of Southampton

Prof Phil LUTHERT, UCL Institute of Ophthalmology

Dr Tom MACGILLIVRAY, University of Edinburgh

Dr Sarah MACKIE, University of Leeds

Dr Bernadette MCGUINNESS, Queen’s University Belfast

Dr Gareth MCKAY, Queen’s University Belfast

Dr Martin MCKIBBIN, Leeds Teaching Hospitals NHS Trust

Prof Tony MOORE, UCL Institute of Ophthalmology

Prof James MORGAN, Cardiff University

Prof Richard ORAM, University of Exeter

Dr Eoin O’SULLIVAN, King’s College Hospital

Prof Chris OWEN, St George’s, University of London

Dr Praveen PATEL, Moorfields Eye Hospital

Dr Euan PATERSON, Queen’s University Belfast

Dr Tunde PETO, Queen’s University Belfast

Dr Axel PETZOLD, UCL Institute of Neurology

Dr Nikolas PONTIKOS, UCL Institute of Ophthalmology

Prof Jugnoo RAHI, UCL Institute of Child Health

Prof Alicja RUDNICKA, St George’s, University of London

Prof Naveed SATTAR, University of Glasgow

Dr Jay SELF, University of Southampton

Dr Panagiotis SERGOUNIOTIS, The University of Manchester

Prof Sobha SIVAPRASAD, Moorfields Eye Hospital

Prof David STEEL, Newcastle University

Ms Irene STRATTON, Gloucestershire Hospitals NHS Foundation Trust

Dr Nicholas STROUTHIDIS, Moorfields Eye Hospital

Prof Cathie SUDLOW, University of Edinburgh

Dr Zihan SUN, UCL Institute of Ophthalmology

Dr Robyn TAPP, St George’s, University of London

Dr Dhanes THOMAS, Moorfields Eye Hospital

Prof Emanuele TRUCCO, University of Dundee

Prof Adnan TUFAIL, Moorfields Eye Hospital

Dr Ananth VISWANATHAN, Moorfields Eye Hospital

Dr Veronique VITART, University of Edinburgh

Dr Mike WEEDON, University of Exeter

Dr Katie WILLIAMS, King’s College London

Prof Cathy WILLIAMS, University of Bristol

Prof Jayne WOODSIDE, Queen’s University Belfast

Dr Max YATES, University of East Anglia

Dr Jennifer YIP, University of Cambridge

Dr Yalin ZHENG, University of Liverpool

