## Supplementary Materials for "Artificial intelligence enabled retinal vasculometry for prediction of circulatory mortality, myocardial infarction and stroke"

#### UK Biobank biomedical examination

Baseline assessments were carried out 2006-2010, in 22 UK recruitment centres, in 502,682 adults aged 40-69 years.[1] Study participants had a detailed examination (including anthropometry, blood pressure, urine and venous blood sampling) and self-completed questionnaire about health (including information on pre-existing CVD, self-reported heart attack, stroke, angina, type 2 diabetes, and other medical conditions), and lifestyle (with a particular focus on dietary habits and smoking status) as well as medication usage (including lipid lowering, antihypertensives and insulin). Weight and height, were measured in participants after removal of heavy clothing and without shoes. Weight was measured using digital scales (Tanita BC-418MA, Tanita UK Ltd, Middlesex, UK) and height with a stadiometer (Seca 202, Seca, Birmingham, UK). Seated blood pressure was measured twice 1 minute apart using an automated blood pressure monitor (Omron HEM-7015IT, Omron Electronics Ltd, Milton Keynes, UK); the mean of both measures was used. A non-fasting venous blood sample was collected; details of the analytic measures have been published previously.[2] Blood samples were processed and analysed by a single laboratory between 2014-2017, and included serum total cholesterol, and HDL-cholesterol;[3] LDL-cholesterol was calculated using the Fredrickson–Friedewald equation,[4] except in 10,884 patients where triglycerides were >400 mg/dL (2.2%) where a direct measure was used.[3]

UK Biobank eye examination occurred at baseline in a subset of participants[5] from December 2009 to July 2010 towards the latter end of recruitment in 6 UK Biobank centres. Participants attended for repeat assessment 1 to 5 years after recruitment and ocular assessments in this latter phase (August 2012-June 2013) were largely from individuals that had not undergone an ocular assessment on entry into UK Biobank. Both phases included visual acuity, autorefraction, intraocular pressure and corneal biomechanics.[5] Digital fundus photography and spectral domain OCT images were taken using the Topcon 3D-OCT 1000 Mark 2. Non-mydriatic 45° digital colour images, centred on the fovea were captured from 68,550 participants in the first phase and 19,502 from the second phase. Overlap with baseline ocular assessment was minimal.

#### EPIC-Norfolk biomedical examination at 3rd Health Check

Between 2004 and 2011 8,623 participants took part in the third health check. Weight and height, were measured with participants in light clothing without shoes. Weight was measured to the nearest 0.1 kg using regularly calibrated digital scales (Tanita TBF-300, Tanita UK Ltd, Middlesex, UK) and height to the last complete 0.1 cm using a stadiometer (Chasmors, UK). Seated blood pressure was measured twice using an automated blood pressure monitor (Accutorr PlusTM, Datascope Patient Monitoring, Huntington,

UK); the mean of both measures was used. A non-fasting venous blood sample was collected; details of the analytic measures have been published previously.[6] Serum total cholesterol and HDL-cholesterol were measured using an auto-analyser (RA 1000 Technicon, Bayer Diagnostics, Basingstoke, UK); LDL-cholesterol was calculated using the Fredrickson–Friedewald equation.[4]

EPIC- Norfolk eye examination. Ophthalmic tests included measurement of vision, visual acuity (LogMAR acuity), and closed field auto-refraction (Humphrey model 500, Humphrey Instruments, San Leandro, California, USA), which was used to estimate axial length. Macular centred 45° digital fundus photographs were taken using a TRC-NW6S non-mydriatic retinal camera and IMAGEnet Telemedicine System (Topcon Corporation, Tokyo, Japan) with a 10 megapixel Nikon D80 camera (Nikon Corporation, Tokyo, Japan) without pharmacological dilation of the pupil.

#### Health outcomes

The primary outcome was circulatory mortality as defined using International Classification of Diseases (ICD-10 codes I00-I99 and ICD9 390-459) coded death registry data from the Office for National Statistics and the Health and Social Care Information Centre (now NHS Digital) for England and Wales, and the Information Services Department for Scotland, provided information on date and cause(s) of death to 31st January 2018 for UK Biobank and 31st March 2018 for EPIC-Norfolk. Incident MI and stroke events after retinal image capture were based on medical records linkage with hospital diagnoses of non-fatal events, supplemented with participant health and lifestyle questionnaire data from repeat surveys in UK Biobank and EPIC-Norfolk (2012-2018). ICD-10 codes I21-I25 (or ICD-9 codes 410, 411,412 429.79) were used for fatal and non-fatal MI; and ICD-10 codes I60,61,63,64 (or ICD-9 codes 430, 431,434,436) for ischaemic and haemorrhagic stroke.

#### Statistical Analysis

##### Development of circulatory mortality models in UK Biobank

Statistical analyses were carried out using STATA software (version 16, StataCorp LP, College Station, TX). Retinal vessel widths and area showed normal distributions, tortuosity required log-transformation and within-vessel-width-variance required inverse square-root transformation to normalize distributions. Throughout models were developed in UK Biobank for men and women separately, and externally validated in EPIC-Norfolk. We hypothesized that retinal vessel characteristics in relation to disease incidence, might be modified by age, smoking status, presence of CVD/diabetes and use of BP lowering medications. Hence, two-way interactions between retinal vasculometry and age, smoking status and self-reported use of blood pressure medication, prevalent diabetes and CVD were first examined in mutually adjusted Cox proportional hazard[7] models for circulatory mortality. Interaction terms with p values <0.2 were then included along with main effects in Cox regressions models using backward elimination (p value set to 0.1).

Bootstrapping with 100 replications was used for internal validation to adjust model performance measures for optimism, including Harrel's C-statistic for discrimination, $R^2$ statistic (representing a measure of explained variation) and calibration slope (where a slope of 1.0 is ideal).[8] The model from the bootstrapped sample was applied to the bootstrapped sample to estimate *apparent performance* and to the original dataset to test *model performance*. Optimism was estimated within each bootstrapped sample as the difference in performance parameters (C-statistic, $R^2$ and calibration slope) between *model performance vs apparent performance*. The overall (average) optimism across all bootstrapped samples was determined to adjust measures of model performance (C-statistic, $R^2$ and calibration slope).

##### External validation of circulatory mortality models in EPIC-Norfolk cohort

The original beta coefficients from the prognostic models were adjusted for shrinkage to allow for over-fitting using the calibration slopes adjusted for optimism from the bootstrapped sampling. The adjusted linear predictor was then applied to the EPIC-Norfolk cohort and C-statistic, $R^2$ and calibration slope estimated. Calibration plots of the observed vs expected event probability by octiles of predicted risk of an event were calibrated to the average 5-year baseline survival in the EPIC-Norfolk cohort.

##### Framingham Risk Scores for stroke and MI in UK Biobank and EPIC-Norfolk cohorts

Framingham risk scores (FRS) for incident fatal and non-fatal stroke[9] and MI[10] were applied to UK Biobank and EPIC-Norfolk cohorts and recalibrated to baseline survival function within each cohort. Following FRS criteria, participants reporting use of cholesterol lowering medications, diabetes or missing data on total or HDL cholesterol were excluded from all MI analyses.[10] Those reporting a history of heart attack or stroke or those with a date of event stroke or MI prior to retinal image capture were excluded from the corresponding prognostic modelling for that outcome. FRS models were also extended to include retinal vasculometry. Model development and validation followed a similar approach as described for circulatory mortality.

##### Retinal vasculometry models for stroke and MI in UK Biobank and EPIC-Norfolk cohort

Alternative models for incident fatal and non-fatal stroke and MI using age, smoking status, medical history (self-reported history of heart attack, stroke or diabetes and use of blood pressure lowering medications) and retinal vasculometry only were developed in UK Biobank following the same approach as for circulatory mortality. A medical history of MI did not preclude inclusion in models for incident stroke events and vice-versa. Participants reporting diabetes or use of blood pressure lowering medications were included in stroke analyses. Participants with missing data on smoking status or self-report on medications for lowering blood pressure or lipids, or those that preferred not to report a history of heart attack or stroke were excluded from all FRS analyses (UK Biobank n= 1182 (1.8%); EPIC-Norfolk n=93 (1.6%)).

Prognostic models using retinal vasculometry included up to 26 candidate predictors in men and up to 28 in women, in the stepwise procedure based on inclusion of main effects and interactions with retinal vasculometry with $p<0.2$. A maximum of 16 predictors were identified by the stepwise procedure with $p<0.1$ in any single model. Retinal vasculometry measures excluded by the stepwise procedure were re-inserted back into the model to check whether they became statistically significant. Fractional polynomial models were used to examine presence of non-linear associations but none were identified.

##### Sensitivity analyses

Sensitivity analyses restricted the entire model development and validation to the white ethnic group to check for systematic differences in model performance. With the EPIC-Norfolk cohort having a relatively smaller number of incident events, we assessed the external validation of models to a broader spectrum of incident cerebrovascular disease (ICD10 I60-69; ICD 9 430-438) and incident ischaemic heart disease (ICD10 I20-I25; ICD9 410-414).

#### Sample size considerations

Prediction models considered the following variables: retinal vessel width, tortuosity, area, width variance [arteriolar and venular], age, sex, smoking status [current, former and never], blood pressure, serum lipids [total and HDL cholesterol] Framingham risk scores, history of diabetes / stroke / heart attack, use of blood pressure lowering medications plus significant two-way interactions with retinal vasculometry (described above). This yielded between 26 to 28 candidate predictor parameters for consideration in the stepwise regression procedure. With 65,000 UK Biobank participants, 327 circulatory deaths, 446 incident strokes and 393 incident MI events provided sufficient sample size to ensure model shrinkage factor (to allow for over-fitting) was in the region of 0.9 and that absolute differences in model's apparent vs an adjusted $R^2$ (hypothesized to be ~0.2), was approximately 0.1.[11] UK Biobank provided an unprecedented sample size in terms of retinal imaging on a population based sample. It encompassed a wide range of patient characteristics for model development and it has been shown that risk factor associations in the UK Biobank seem to be generalisable.[12]

#### Ethics, governance and consent

The UK Biobank and EPIC-Norfolk studies were carried out following the principles of the Declaration of Helsinki and the Research Governance Framework for Health and Social Care. The UK Biobank study was approved by the North West Multi-Centre Research Ethics Committee (11/NW/03820). All participants gave written, informed consent.

The EPIC-Norfolk study was approved by the Norfolk Local Research Ethics Committee (05/Q0101/191) and East Norfolk and Waveney NHS Research Governance Committee (2005EC07L). All participants gave written, informed consent.

The data reported in this article are available via application to the UK Biobank to other researchers for purposes of reproducing the results or replicating the procedure.

### Supplemental Figures

#### Figure S1 Participant flow chart in UK Biobank and EPIC cohorts

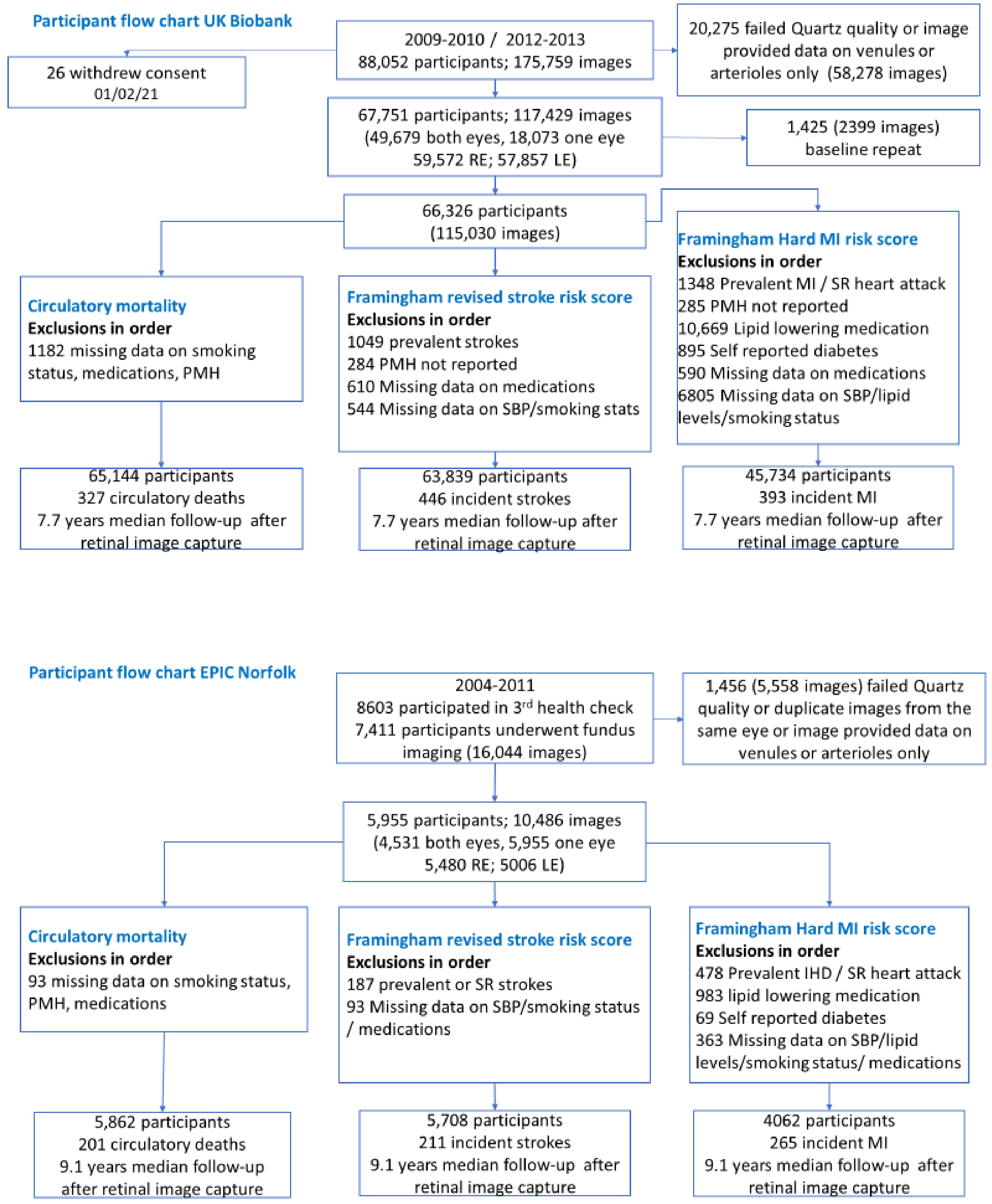

SR= self-reported; PMH previous medical history; SBP systolic blood pressure

Figure S2 Observed risk of incident stroke at 5 years by deciles of predicted risk in UK Biobank and octiles of predicted risk in EPIC-Norfolk

**UK Biobank Men** **UK Biobank Women** **EPIC-Norfolk Men** **EPIC- Norfolk Women**

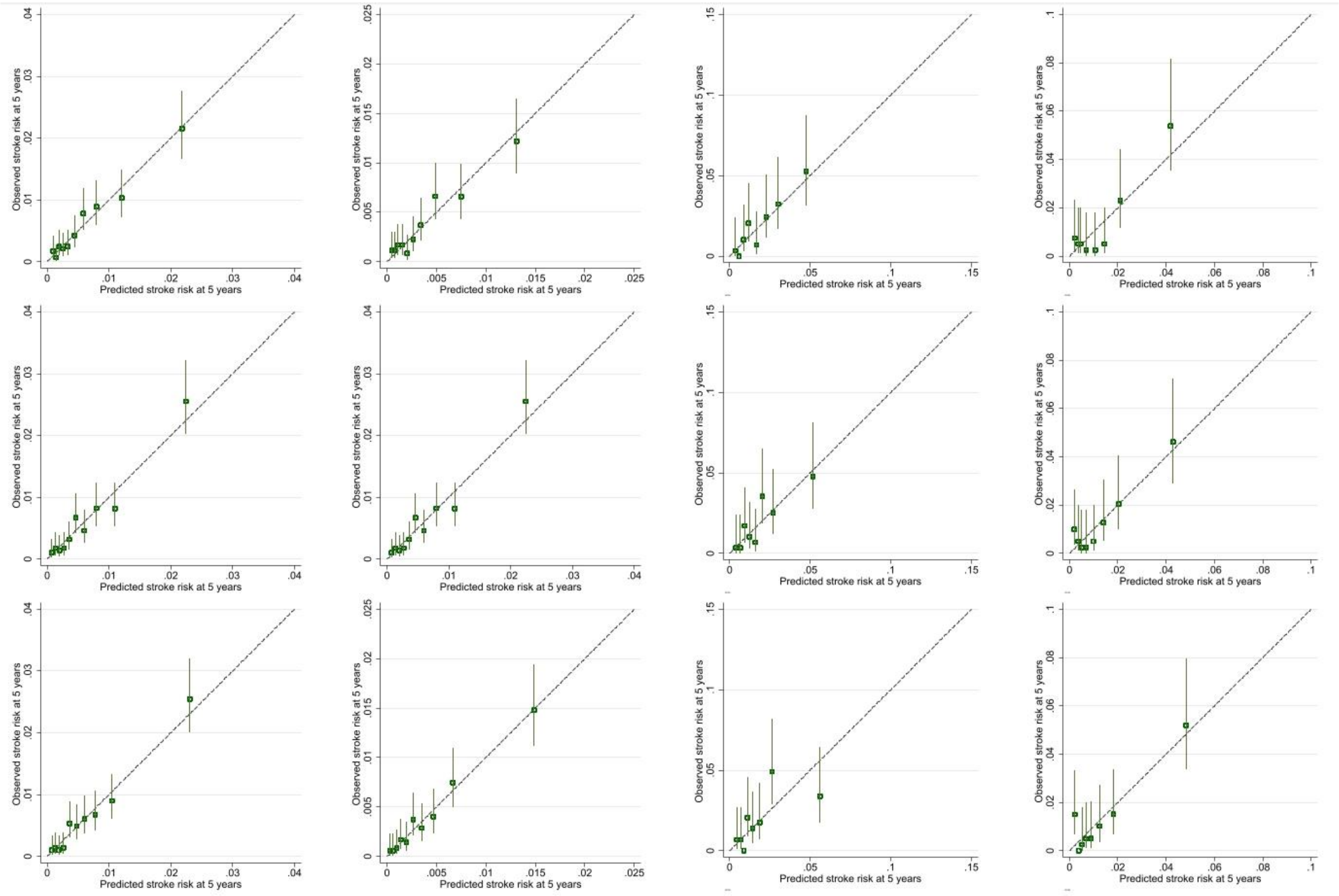

***Figure S2 footnote:***

*Top row: revised Framingham stroke risk score (after recalibration for baseline survival within each cohort)*

*Middle row: prediction model based on revised Framingham stroke risk score plus retinal vasculometry*

*Bottom row: prediction model based on retinal vasculometry, age, smoking and medical history*

*Vertical lines around symbols are 95% confidence intervals. Dotted line represents perfect calibration.*

*Incident stroke codes: ICD10: I60,I61,I63,I64, ICD9: 430,431,434,436*

*The scale of the vertical and horizontal axes is a probability e.g., 0.1 equates to a 10% risk of event by 5 years.*

Figure S3 Observed risk of confirmed MI at 5 years by deciles of predicted risk in UK Biobank and octiles of predicted risk in EPIC-Norfolk

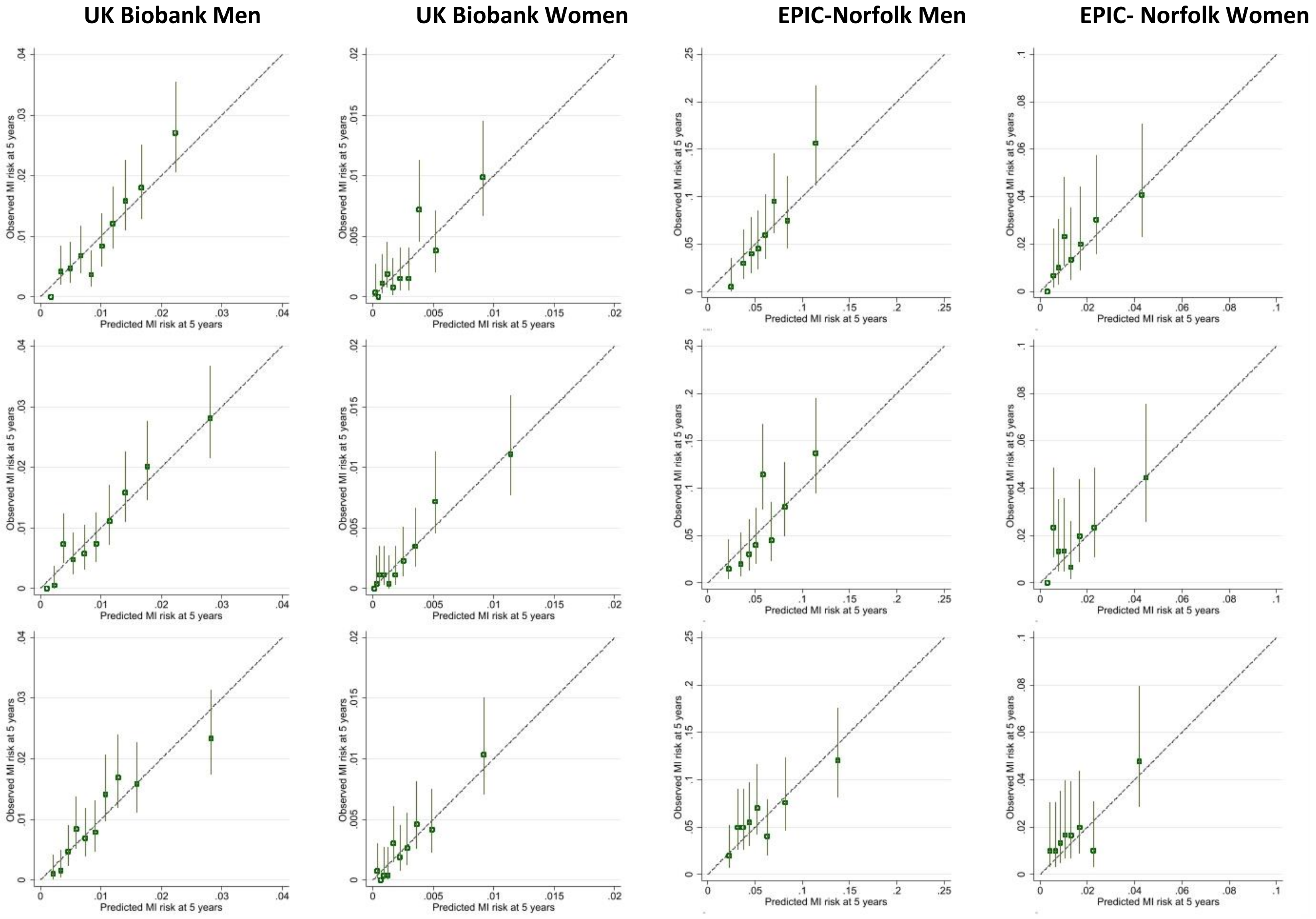

***Figure S3 footnote***:

*Top row: Framingham risk score for confirmed MI (after recalibration for baseline survival within each cohort)*

*Middle row: prediction model based on Framingham risk score for confirmed MI plus retinal vasculometry*

*Bottom row: prediction model based on retinal vasculometry, age, smoking and medical history*

*Vertical lines around symbols are 95% confidence intervals. Dotted line represents perfect calibration.*

*Incident MI codes: ICD10: I21-I25, ICD9: 410,411,412,429.79*

*The scale of the vertical and horizontal axes is a probability e.g., 0.1 equates to a 10% risk of event by 5 years.*

Figure S4 Observed risk of incident cerebrovascular disease at 5 years by eighths of predicted risk in EPIC-Norfolk cohort

**EPIC-Norfolk Men** **EPIC- Norfolk Women**

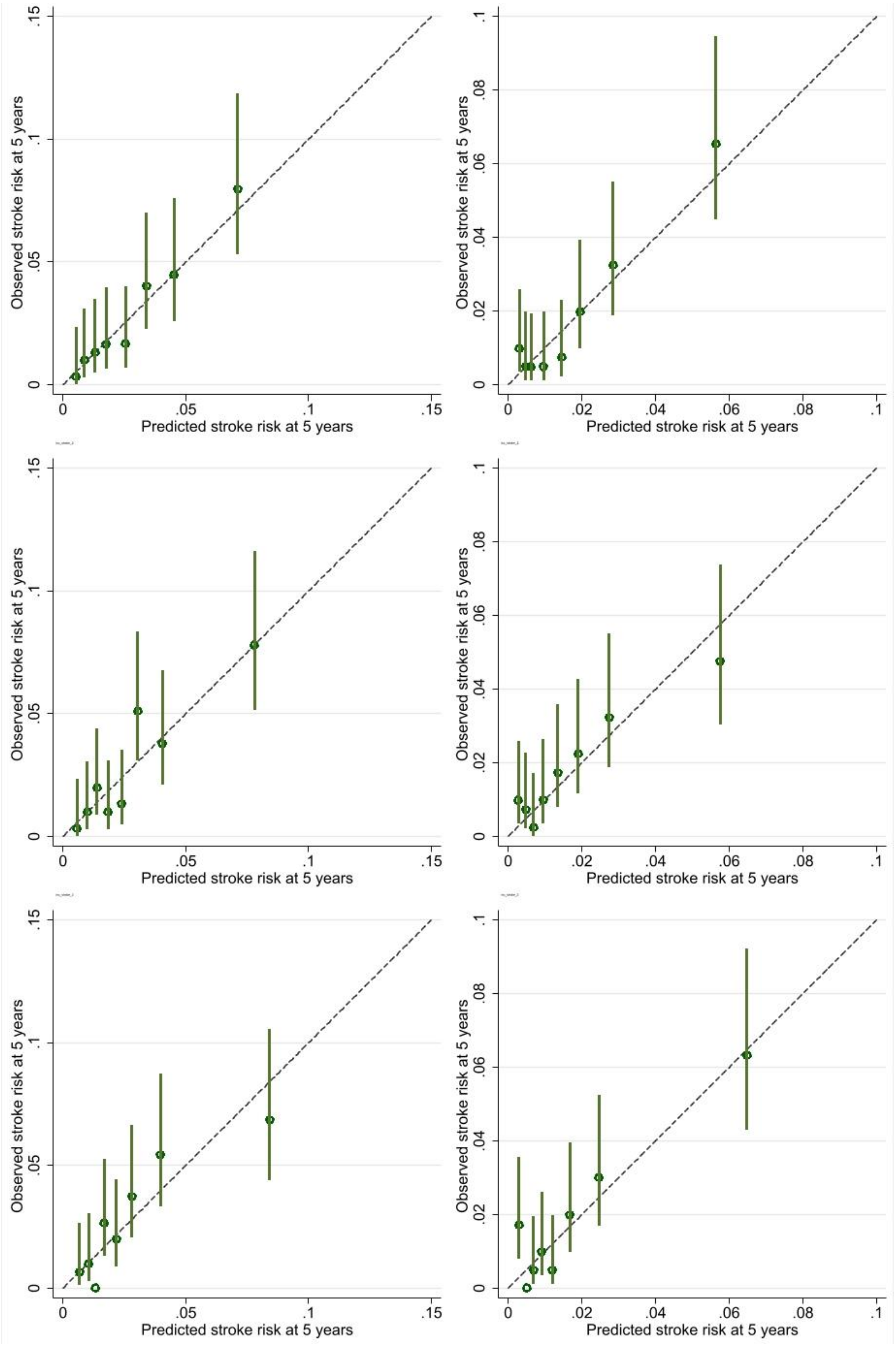

***Figure S4 footnote***:

*Incident cerebrovascular disease ICD9 430-438; ICD10 I60-I69*

*Top row: revised Framingham stroke risk score (after recalibration for baseline survival in EPIC-Norfolk)*

*Middle row: prediction model based on revised Framingham stroke risk score plus retinal vasculometry*

*Bottom row: prediction model based on retinal vasculometry, age, smoking and medical history*

*Vertical lines around symbols are 95% confidence intervals. Dotted line represents perfect calibration.*

Figure S5 Observed risk of ischaemic heart disease at 5 years by eighths of predicted risk for in EPIC-Norfolk cohort

**EPIC-Norfolk Men** **EPIC- Norfolk Women**

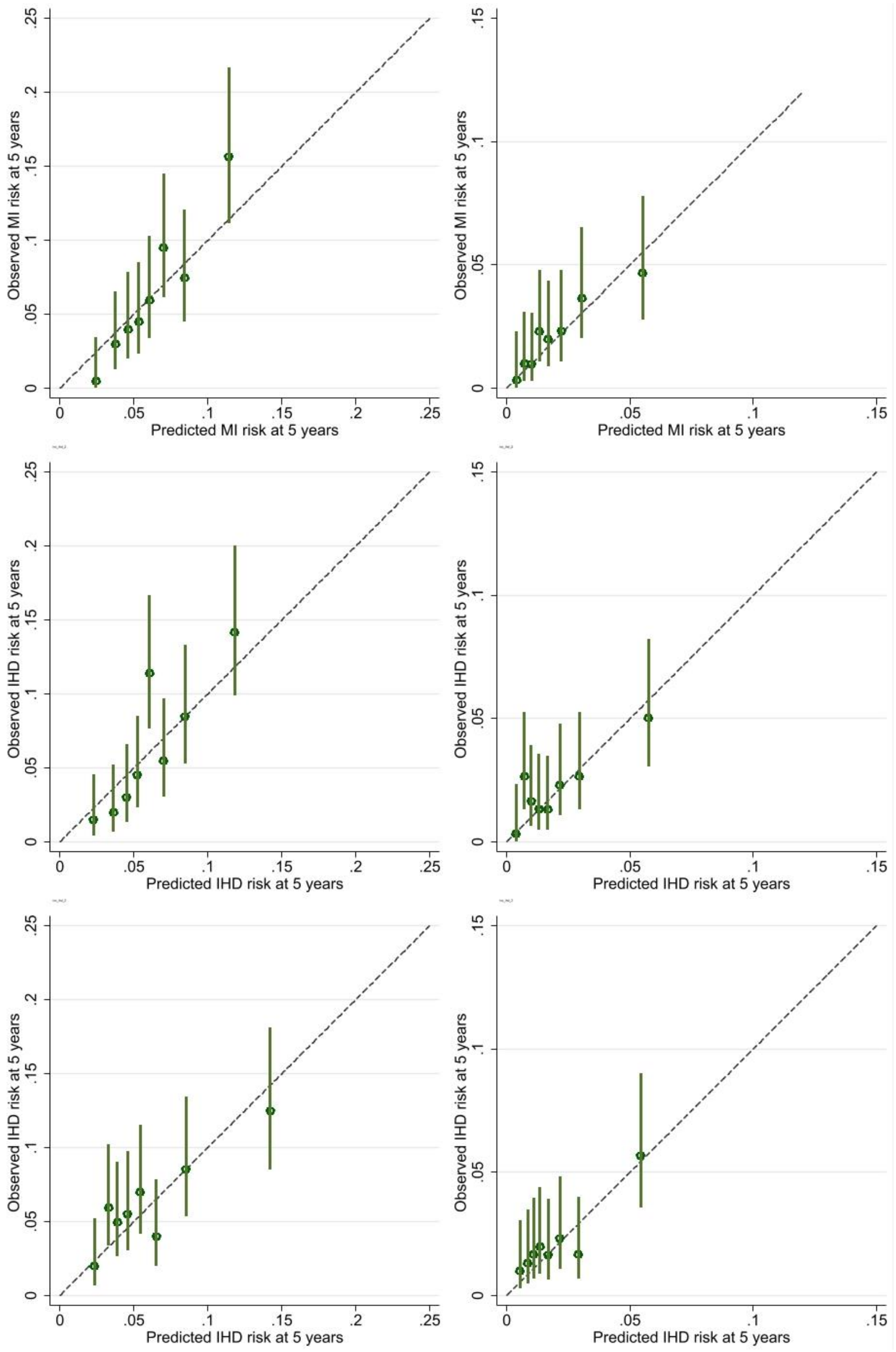

***Figure S5 footnote***:

*Ischaemic heart disease codes ICD9 410-414; ICD10 I20-I25*

*Top row : Framingham risk score for confirmed MI (after recalibration for baseline survival in EPIC-Norfolk)*

*Middle row: prediction model based on Framingham risk for confirmed MI score plus retinal vasculometry*

*Bottom row: prediction model based on retinal vasculometry, age, smoking and medical history*

*Vertical lines around symbols are 95% confidence intervals. Dotted line represents perfect calibration.*

### Supplementary Tables S1 to S6

Table S1 Model diagnostics (with 95% confidence intervals) from internal validation of circulatory mortality in UK Biobank (2009-2018). External validation in EPIC- Norfolk cohort using biomedical data from the third health check (2004-2011) with circulatory mortality (ICD-10 codes I00-I99) as the health outcome (2004-2018)

| **Model** | **Apparent performance** | **Test performance** | **Average Optimism** | **Optimism corrected** |
|---|---|---|---|---|
| **UK Biobank Men** | | | | |
| **Age, smoking, medical history and retinal vasculometry** | | | | No. events = 114 |
| Calibration Slope | 1.000 (0.887, 1.113) | 0.913 (0.834, 0.992) | 0.087 | 0.913 (0.800, 1.026) |
| C-statistic | 0.771 (0.741, 0.800) | 0.763 (0.752, 0.773) | 0.021 | 0.749 (0.720, 0.779) |
| $R^2$ | 0.418 (0.359, 0.476) | 0.400 (0.384, 0.417) | 0.049 | 0.369 (0.310, 0.427) |
| **UK Biobank Women** | | | | |
| **Age, smoking, medical history and retinal vasculometry** | | | | No. events = 87 |
| Calibration Slope | 1.000 (0.875, 1.125) | 0.857 (0.708, 1.006) | 0.143 | 0.857 (0.732, 0.982) |
| C-statistic | 0.799 (0.753, 0.846) | 0.787 (0.766, 0.808) | 0.036 | 0.763 (0.717, 0.810) |
| $R^2$ | 0.522 (0.448, 0.597) | 0.488 (0.449, 0.526) | 0.079 | 0.443 (0.369, 0.518) |

Table S2 Final multivariable models based on retinal vasculometry, age, smoking status and medical history for circulatory mortality, incident stroke, incident myocardial infarctions (MI) in MEN. For each model the mean (standard deviation) of the linear predictor is also given

| Model | Hazard ratio (95%CI) | β coefficients |
|---|---|---|
| **Age, smoking, medical history and retinal vasculometry** | **Circulatory Mortality** | |
| Age | 1.08 (1.05, 1.10) | 0.07356 |
| Taking BP lowering Medication | 1.59 (1.18, 2.13) | 0.46127 |
| Previous MI | 3.87 (2.75, 5.45) | 1.35365 |
| Previous stroke | 2.35 (1.44, 3.84) | 0.85541 |
| Diabetes | 2.25 (1.61, 3.15) | 0.81162 |
| Current smoker | 2.33 (1.55, 3.48) | 0.84374 |
| Arteriolar InvSD | 0.93 (0.86, 1.01) | -0.07171 |
| Venular InvSD | 1.07 (1.00, 1.14) | 0.06708 |
| Age # arteriolar width | 1.00 (1.00, 1.00) | 0.00194 |
| Venular tortuosity if occasional smoker | 0.12 (0.05, 0.30) | -2.13421 |
| Venular width if non-smoker | 1.02 (1.00, 1.04) | 0.01895 |
| Arteriolar width if non-smoker | 0.96 (0.93, 0.99) | -0.04003 |
| Mean (SD) of linear predictor | 0.4066 (0.9699) | |
| **Age, smoking, medical history + retinal vasculometry** | **Incident stroke** | |
| Age | 1.10 (1.08, 1.13) | 0.09860 |
| Current smoker | 3.10 (2.02, 4.76) | 1.13170 |
| Diabetes | 1.78 (1.26, 2.53) | 0.57847 |
| History of CVD | 2.05 (1.32, 3.18) | 0.71631 |
| Venular width | 0.99 (0.98, 1.00) | -0.01136 |
| Venular tortuosity if history of CVD | 0.40 (0.17, 0.94) | -0.90743 |
| Arteriolar tortuosity if taking BP lowering medication | 0.68 (0.46, 1.01) | -0.37987 |
| Venular width if taking BP lowering medication | 1.03 (1.01, 1.05) | 0.02592 |
| Arteriolar width if previous smoker | 0.97 (0.95, 1.00) | -0.02561 |
| Arteriolar tortuosity if occasional smoker | 0.38 (0.12, 1.18) | -0.95683 |
| Venular tortuosity width if previous smoker | 1.74 (1.02, 2.98) | 0.55405 |

| Model | Hazard ratio (95%CI) | β coefficients |
|---|---|---|
| Venular tortuosity if current smoker | 4.37 (1.74, 11.01) | 1.47586 |
| Mean (SD) of linear predictor | 0.2347 (0.9762) | |
| **Age, smoking, medical history + retinal vasculometry** | **Incident MI** | |
| Age | 1.07 (1.05, 1.09) | 0.06901 |
| History of CVD | 2.39 (1.22, 4.69) | 0.87216 |
| Taking BP lowering Medication | 1.45 (1.07, 1.97) | 0.37401 |
| Current smoker | 3.19 (2.29, 4.45) | 1.16050 |
| Arteriolar width | 0.98 (0.96, 1.00) | -0.02412 |
| Age # arteriolar area | 1.02 (1.00, 1.04) | 0.02039 |
| Arteriolar width if non-smoker | 1.03 (1.00, 1.06) | 0.02702 |
| Venular width if occasional smoker | 0.92 (0.88, 0.97) | -0.07865 |
| Venular InvSD if previous smoker | 0.93 (0.87, 0.99) | -0.07380 |
| Mean (SD) of linear predictor | 0.1050 (0.7534) | |

FRS = Framingham risk score for outcomes as defined in methods

Age is in years centred to 55 years, SBP systolic blood pressure in mmHg

Arteriolar and venular widths are in microns centred to 85 microns and 100 microns respectively

Arteriolar and venular vessel area are in $mm^2$

* InvSD is the transformed segment-width-variance values x100 (a unit increase equates to approximately 0.5 standard deviations)

### indicates interaction term between continuous variables

All regression coefficients are per unit increase in the predictors

Table S3: Final multivariable models based on retinal vasculometry, age, smoking status and medical history for circulatory mortality, incident stroke, myocardial infarctions (MI) and in WOMEN For each model the mean (standard deviation) of the linear predictor is also given

| **Model** | **Hazard ratio (95%CI)** | **β coefficients** |
|---|---|---|
| | **Circulatory mortality** | |
| Age | 1.108 (1.071, 1.147) | 0.10285 |
| Taking BP lowering medication | 1.823 (1.166, 2.849) | 0.60032 |
| Diabetes | 3.754 (2.211, 6.375) | 1.32294 |
| Occasional smoker | 1.000 (0.000, 0.000) | 0.00000 |
| Current smoker | 2.755 (1.603, 4.736) | 1.01350 |
| Arteriolar area | 0.172 (0.072, 0.410) | -1.76009 |
| Venular area | 1.605 (1.092, 2.358) | 0.47298 |
| Venular InvSD | 0.676 (0.587, 0.779) | -0.39135 |
| Venular area if not taking BP lowering medication | 0.492 (0.305, 0.793) | -0.70972 |
| Arteriolar area if non-smoker | 2.638 (1.597, 4.359) | 0.97007 |
| Venular InvSD and no history of MI | 1.419 (1.222, 1.650) | 0.35028 |
| Arteriolar width and no history of stroke | 1.026 (0.999, 1.054) | 0.02603 |
| Arteriolar area and no history of stroke | 3.205 (1.354, 7.582) | 1.16461 |
| Venular width if non-smoker | 0.975 (0.954, 0.997) | -0.02489 |
| Venular tortuosity if previous-smoker | 6.168 (2.729, 13.941) | 1.81938 |
| Arteriolar width if previous-smoker | 0.950 (0.909, 0.992) | -0.05179 |
| Mean (SD) of linear predictor | 0.4356 (1.0503) | |
| | **Incident stroke** | |
| Age | 1.103 (1.077, 1.130) | 0.09808 |
| Taking BP lowering medication | 1.580 (1.141, 2.189) | 0.45746 |
| History of CVD | 2.341 (1.413, 3.879) | 0.85059 |
| Diabetes | 3.151 (2.011, 4.939) | 1.14778 |
| Venular area | 1.786 (1.111, 2.871) | 0.58011 |
| Arteriolar area | 1.707 (1.037, 2.808) | 0.53465 |
| Arteriolar tortuosity | 1.572 (1.057, 2.338) | 0.45247 |
| Venular tortuosity | 1.410 (0.903, 2.202) | 0.34357 |
| Age # arteriolar tortuosity | 0.952 (0.913, 0.993) | -0.04913 |
| Venular area if not taking BP lowering medication | 0.697 (0.481, 1.012) | -0.36031 |
| Arteriolar area if do not have diabetes | 0.453 (0.265, 0.773) | -0.79244 |

| Model | Hazard ratio (95%CI) | β coefficients |
| --- | --- | --- |
| Venular area if do not have diabetes | 0.615 (0.365, 1.036) | -0.48565 |
| Venular tortuosity if ex-smoker | 2.764 (1.357, 5.628) | 1.01650 |
| Venular width if occasional smoker | 1.059 (1.035, 1.085) | 0.05766 |
| Mean (SD) of linear predictor | 0.0589 (1.1128) | |
| | **Incident MI** | |
| Age | 1.093 (1.063, 1.125) | 0.08936 |
| Taking BP lowering medication | 1.637 (1.045, 2.564) | 0.49259 |
| Current smoker | 3.785 (2.214, 6.468) | 1.33094 |
| Venular InvSD | 1.077 (1.009, 1.149) | 0.07400 |
| Venular tortuosity if non-smoker | 1.929 (1.007, 3.695) | 0.65682 |
| Arteriolar area if non-smoker | 0.667 (0.473, 0.940) | -0.40534 |
| Venular area if non-smoker | 0.750 (0.561, 1.004) | -0.28704 |
| Mean (SD) of linear predictor | 0.0394 (0.9406) | |

FRS = Framingham risk score for outcomes as defined in the methods

Age is in years, SBP systolic blood pressure in mmHg

Arteriolar and venular widths are in microns centred to 85 microns and 100 microns respectively

Arteriolar and venular vessel area are in $mm^2$

*InvSD is the transformed segment-width-variance values x100 (a unit increase equates to approximately 0.5 standard deviations)

### indicates interaction term between continuous variables

All regression coefficients are per unit increase in the predictors

Table S4 Model diagnostics (with 95% confidence intervals) for incident stroke (after retinal image capture) in UK Biobank (2009-2018) as defined in the methods. External validation in EPIC- Norfolk cohort using biomedical data from the third health check (2004-2011) with **all** incident cerebrovascular disease (ICD10 I60-I69) as the health outcome (2004-2018)

| Model | Apparent performance | Test performance | Average Optimism | Optimism corrected | External validation in EPIC-Norfolk |
|---|---|---|---|---|---|
| **Revised FRS stroke** | | **UK Biobank Men** | | | No. events = 176 |
| Calibration Slope | - | - | - | - | 1.019 (0.810, 1.227) |
| C-statistic | - | - | - | - | 0.711 (0.672, 0.749) |
| $R^2$ | - | - | - | - | 0.273 (0.196, 0.350) |
| **Revised FRS stroke + retinal vasculometry** | | | | | |
| Calibration Slope | 1.000 (0.869, 1.131) | 0.911 (0.790, 1.032) | 0.089 | 0.911 (0.780, 1.042) | 0.911 (0.722, 1.100) |
| C-statistic | 0.749 (0.719, 0.780) | 0.742 (0.733, 0.751) | 0.018 | 0.731 (0.701, 0.762) | 0.698 (0.658, 0.739) |
| $R^2$ | 0.359 (0.299, 0.419) | 0.342 (0.323, 0.360) | 0.042 | 0.317 (0.257, 0.377) | 0.248 (0.170, 0.325) |
| **Age, smoking, medical history + retinal vasculometry** | | | | | |
| Calibration Slope | 1.000 (0.871, 1.129) | 0.896 (0.772, 1.019) | 0.104 | 0.896 (0.767, 1.025) | 0.910 (0.736, 1.084) |
| C-statistic | 0.751 (0.721, 0.781) | 0.737 (0.727, 0.747) | 0.022 | 0.729 (0.699, 0.759) | 0.711 (0.672, 0.750) |
| $R^2$ | 0.365 (0.306, 0.425) | 0.330 (0.311, 0.348) | 0.050 | 0.315 (0.256, 0.375) | 0.262 (0.186, 0.337) |
| **Revised FRS stroke** | | **UK Biobank Women** | | | No. events = 190 |
| Calibration Slope | - | - | - | - | 1.079 (0.915, 1.242) |
| C-statistic | - | - | - | - | 0.758 (0.723, 0.794) |
| $R^2$ | - | - | - | - | 0.365 (0.296, 0.435) |
| **Revised FRS stroke + retinal vasculometry** | | | | | |

| Model | Apparent performance | Test performance | Average Optimism | Optimism corrected | External validation in EPIC-Norfolk |
|---|---|---|---|---|---|
| Calibration Slope | 1.000 (0.862, 1.138) | 0.858 (0.656, 1.061) | 0.142 | 0.858 (0.720, 0.996) | 0.923 (0.770, 1.076) |
| C-statistic | 0.771 (0.740, 0.803) | 0.762 (0.752, 0.773) | 0.021 | 0.750 (0.719, 0.782) | 0.731 (0.694, 0.768) |
| $R^2$ | 0.388 (0.323, 0.452) | 0.370 (0.351, 0.390) | 0.051 | 0.337 (0.272, 0.401) | 0.314 (0.242, 0.387) |
| **Age, smoking, medical history + retinal vasculometry** | | | | | |
| Calibration Slope | 1.000 (0.869, 1.131) | 0.860 (0.665, 1.055) | 0.140 | 0.860 (0.729, 0.991) | 0.840 (0.710, 0.971) |
| C-statistic | 0.776 (0.744, 0.807) | 0.766 (0.754, 0.778) | 0.023 | 0.753 (0.721, 0.784) | 0.734 (0.695, 0.773) |
| $R^2$ | 0.408 (0.345, 0.472) | 0.386 (0.363, 0.409) | 0.056 | 0.352 (0.289, 0.416) | 0.323 (0.251, 0.394) |

*FRS=Framingham risk score*

Table S5 Model diagnostics (with 95% confidence intervals) for incident myocardial infarction (after retinal image capture) in UK Biobank (2009-2018) as defined in the methods. External validation in EPIC- Norfolk cohort using biomedical data from the third health check (2004-2011) with all incident ischaemic heart disease (ICD10 I20-I25) as the health outcome (2004-2018)

| Model | Apparent performance | Test performance | Average Optimism | Optimism corrected | External validation in EPIC |
|---|---|---|---|---|---|
| **FRS for confirmed MI (FRS)** | | **UK Biobank Men** | | | No. events = 173 |
| Calibration Slope | - | - | - | - | 1.562 (1.212, 1.912) |
| C-statistic | - | - | - | - | 0.689 (0.651, 0.727) |
| $R^2$ | - | - | - | - | 0.231 (0.153, 0.308) |
| **FRS + retinal vasculometry** | | | | | |
| Calibration Slope | 1.000 (0.838, 1.162) | 0.887 (0.771, 1.002) | 0.113 | 0.887 (0.725, 1.049) | 1.398 (1.072, 1.723) |
| C-statistic | 0.724 (0.697, 0.751) | 0.719 (0.710, 0.728) | 0.020 | 0.704 (0.677, 0.731) | 0.683 (0.646, 0.721) |
| $R^2$ | 0.270 (0.210, 0.330) | 0.259 (0.245, 0.273) | 0.043 | 0.227 (0.167, 0.287) | 0.212 (0.136, 0.288) |
| **Age, smoking, medical history + retinal vasculometry** | | | | | |
| Calibration Slope | 1.000 (0.837, 1.163) | 0.836 (0.715, 0.957) | 0.164 | 0.836 (0.673, 0.999) | 0.910 (0.664, 1.155) |
| C-statistic | 0.704 (0.676, 0.732) | 0.689 (0.677, 0.701) | 0.029 | 0.675 (0.647, 0.703) | 0.641 (0.599, 0.683) |
| $R^2$ | 0.242 (0.182, 0.302) | 0.213 (0.189, 0.236) | 0.064 | 0.178 (0.118, 0.238) | 0.151 (0.079, 0.223) |
| **FRS for confirmed MI (FRS)** | | **UK Biobank Women** | | | No. events = 116 |
| Calibration Slope | - | - | - | - | 0.883 (0.650, 1.116) |
| C-statistic | - | - | - | - | 0.694 (0.649, 0.740) |
| $R^2$ | - | - | - | - | 0.228 (0.135, 0.320) |
| **FRS + retinal vasculometry** | | | | | |

| Model | Apparent performance | Test performance | Average Optimism | Optimism corrected | External validation in EPIC |
|---|---|---|---|---|---|
| Calibration Slope | 1.000 (0.823, 1.177) | 0.849 (0.678, 1.021) | 0.151 | 0.849 (0.672, 1.026) | 0.678 (0.467, 0.890) |
| C-statistic | 0.794 (0.756, 0.831) | 0.786 (0.769, 0.803) | 0.028 | 0.766 (0.728, 0.803) | 0.670 (0.623, 0.717) |
| $R^2$ | 0.420 (0.338, 0.501) | 0.401 (0.371, 0.430) | 0.066 | 0.354 (0.272, 0.435) | 0.167 (0.080, 0.255) |
| **Age, smoking, medical history + retinal vasculometry** | | | | | |
| Calibration Slope | 1.000 (0.787, 1.213) | 0.803 (0.635, 0.970) | 0.197 | 0.803 (0.590, 1.016) | 0.907 (0.661, 1.153) |
| C-statistic | 0.748 (0.708, 0.788) | 0.733 (0.709, 0.757) | 0.039 | 0.709 (0.669, 0.749) | 0.672 (0.620, 0.725) |
| $R^2$ | 0.315 (0.225, 0.405) | 0.292 (0.254, 0.330) | 0.089 | 0.226 (0.136, 0.316) | 0.211 (0.119, 0.303) |

*FRS=Framingham risk score*
